# Invasive Mould Disease in Fatal COVID-19: A Systematic Review of Autopsies

**DOI:** 10.1101/2021.01.13.21249761

**Authors:** Brittany E. Kula, Cornelius J. Clancy, M. Hong Nguyen, Ilan S. Schwartz

**Affiliations:** Division of Infectious Diseases, University of Alberta, Edmonton, Alberta, Canada; Division of Infectious Diseases, University of Pittsburgh, Pittsburgh, Pennsylvania, USA

**Author notes:** Correspondence to: Dr. Ilan S Schwartz MD PhD, Division of Infectious Diseases, Department of Medicine, Faculty of Medicine and Dentistry, University of Alberta, Edmonton, T6G 2G3, Canada. **Funding:** None.

## Abstract

**Background:** Invasive mould disease (IMD) – most commonly pulmonary aspergillosis - is reported to affect up to a third of critically ill COVID-19 patients. Most reported cases are diagnosed with probable/putative COVID-19 associated pulmonary aspergillosis (CAPA) based on a combination of non-specific clinical, radiographic, and mycological findings, but the clinical significance – and whether these cases represent true invasive disease – is unresolved.

**Methods:** We performed a systematic review of autopsy series of decedents with COVID-19 for evidence of IMD. We searched PubMed, Web of Science, OVID (Embase) and MedRxiv for English- or French-language case series published between January 1, 2019 to September 26, 2020. We included series describing lung histology of ≥3 decedents, and authors were contacted for missing information as necessary.

**Findings:** We identified 51 case series describing autopsies of 702 decedents. Individual-level data was available for 430 decedents. The median age was 72 (IQR 61 to 80) years. Diabetes mellitus, pre-existing lung disease, and immunocompromising conditions were reported for 129 (32%), 95 (22%), and 25 (6%) decedents, respectively. The median hospitalization length was 10 (IQR 5-22) days. 51.6% of decedents had received mechanical ventilation for a median of nine (IQR 5-20) days. Treatment included immunomodulation in 60 (most often steroids or tocilizumab) and antifungals in 41 decedents. Eleven decedents (1·6%) had autopsy-confirmed IMD (6 with CAPA, 4 with invasive pulmonary mycosis not specified and 1 with disseminated mucormycosis). Among 173 decedents who received mechanical ventilation, 5 had IMD (2·9%).

**Interpretation:** Autopsy-proven IMD, including CAPA, is uncommon in fatal COVID-19.

**Funding:** This study is unfunded

**Category:** Review

## Background

Coronavirus disease 2019 (COVID-19), caused by the novel severe acute respiratory syndrome Coronavirus 2 (SARS-COV-2), is responsible for a devastating pandemic causing high incidences of critical illness and death among infected persons around the world. In addition to direct viral invasion and downstream immune dysregulation, severe disease can be complicated by secondary infections. Early autopsy studies demonstrated several dominant pathological processes of the lungs including diffuse alveolar damage and thromboembolism.^1^ Secondary infections complicating COVID-19 are reported, but the prevalence of autopsy-proven secondary infections is poorly described.

Secondary fungal infections complicating critical COVID-19 have garnered much attention.^2-5^ Pulmonary aspergillosis, an invasive mould disease (IMD), occurs with increased frequency following other respiratory viral infections such as severe influenza, a complication that has been associated with excess mortality.^6,7^ Similarly, COVID-19-associated pulmonary aspergillosis (CAPA)^8^ has been reported to affect up to 35% of patients with critical COVID-19 requiring intensive care unit (ICU) care;^4,8-10^ in some cohort studies, CAPA has been associated with increased mortality.^11,12^ Risk factors for CAPA appear to include prolonged ICU admission, lymphopenia, and immunosuppression including corticosteroid use.^11,13,14^

Diagnosing IMD in critically ill patients with COVID-19 can be challenging.^15,16^ In practice, given the inherent difficulty in obtaining tissue in critically ill patients, IMD is infrequently histologically proven. Host factors, required for meeting European Organization for Research and Treatment of Cancer and Mycosis Study Group Education and Research Consortium (EORTC/MSGERC) research definitions for probable IMD,^17^ are frequently absent in ICU patients with IPA,^18^ including following severe influenza or COVID-19. Other research definitions for diagnosing CAPA in ICU patients have been proposed and include a combination of non-specific clinical, radiographic, and mycologic criteria.^5,12,15^ Although several ICU cohort studies have suggested high rates of probable or putative CAPA, it is unclear whether these cases represent true tissue-invasive disease. If they do, then IMD would be expected to be a common finding in post-mortem examinations of decedents with fatal COVID-19.

Here we present a systematic review of autopsy studies performed in persons with fatal COVID-19 to determine the prevalence of autopsy-proven tissue-invasive IMD and analyze patient-level data to assess potential risk factors.

## Methods

### Search Strategy and Selection Criteria

This systematic review follows PRISMA reporting guidelines^19^ and the protocol was published as part of a two-study review on PROSPERO (CRD42020204123). We identified possible studies by searching PubMed, Web of Science, OVID (Embase) and MedRxiv for articles published from January 1, 2019 to September 26, 2020. We used the terms “sars cov2” OR “covid” OR “2019 ncov” OR “novel coronavirus” AND “autopsy” OR “post-mortem”. We also reviewed reference lists of eligible publications for additional articles of interest as well as personal databases maintained by the authors. We contacted authors of reports to obtain missing details when required or if the outcome of interest (fungal infections) was not described. We only included articles written in English or French.

Studies met inclusion criteria if they were retrospective or prospective case series of autopsy or post-mortem reports of decedents with confirmed SARS-CoV-2 infection. In order to minimize reporting bias from case reports (either of IMD or competing diagnoses), we only included series with at least three decedents. At minimum, the examination must have included histopathologic investigation of the lungs either through standard or minimally invasive sampling. Studies were excluded if decedents were included or excluded based on diagnoses other than COVID-19, or the presence or absence of complications; or if they were not written in English or French.

All identified titles and abstracts were independently reviewed for potential inclusion criteria by two authors (BEK and ISS) and full texts were obtained for all those aside those deemed ineligible. Full text review was then performed independently by the same two authors (BEK, ISS) for final inclusion. A consensus was reached for all disputes. COVIDENCE was used for abstract management (Veritas Health Innovation, Melbourne, Australia available at www.covidence.org).

In the instance of decedents being included in more than one report, we included the study with the most patient-related variables unless that which contained less individual data had a case of CAPA. If there were more decedents included in the less detailed study, we still extracted data for these remaining patients to add to the total cohort but did not extract any additional patient-level details. When a pre-print went on to be published, we preferentially included publications and excluded the pre-prints as these were not picked up as duplicate studies by our abstract manager.

### Outcomes

The main outcome of interest was the number of decedents with autopsy-proven invasive mould disease (IMD) determined by histopathology showing hyphae invading tissue. When available, we recorded the type of mould as identified in the reports. We recorded the types of autopsies performed as either standard or minimally invasive. Minimally invasive included any autopsy in which multiple lung tissue samples from each lobe were obtained either blindly or through ultrasound guidance; this practice has been applied frequently in COVID-19 to limit occupational exposures at the time of autopsy. We also recorded study country of origin and place of death (hospitalization or community). When possible, individual patient data were also extracted. We collected patient age, comorbidities, length of hospital stay, duration of symptoms, days of mechanical ventilation, and immunomodulatory COVID-19-specific treatment prescribed (i.e. cytokine inhibitors or steroids). We used EORTC/MSGERC definitions of host factors to classify whether decedents were immunocompromised.^17^

### Risk of Bias Assessment

We did not judge the study quality given these are small case series without a target intervention. We attempted to guard against selective reporting by contacting corresponding authors for any manuscript in which the outcome of interest (fungal infection) was not described. We also attempted to mitigate small-study effect by including only those series with three or more cases described.

## Results

We identified 1,070 references from the database searches and 4 from alternative sources; 454 were duplicates, 526 were excluded from title and abstract review and an additional 39 from full text review leaving 51 studies meeting criteria for data extraction (Figure 1). Of these, six were pre-prints and the remainder were published in peer-reviewed journals. All studies were published in 2020.

**Figure 1.**
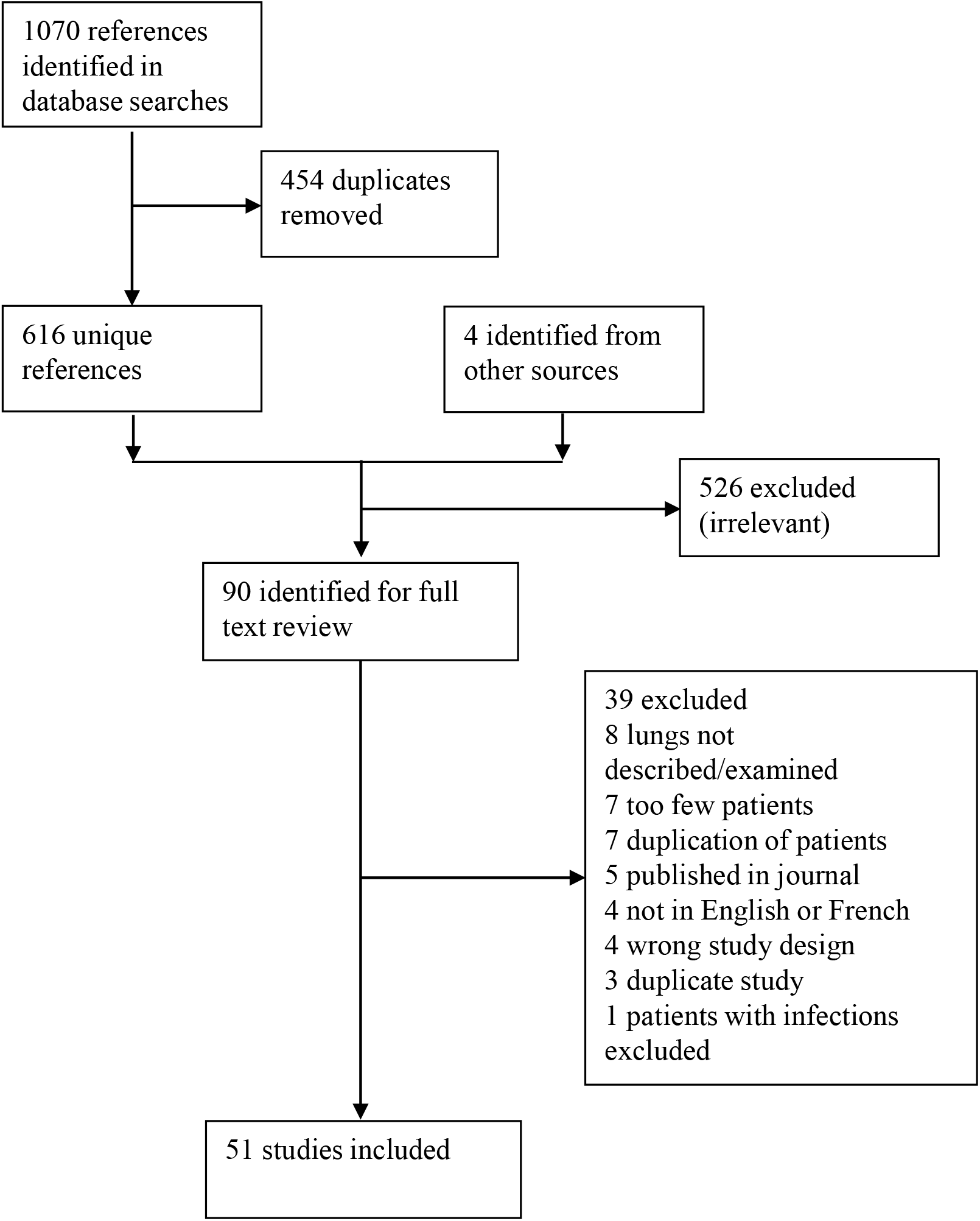

A total of 702 decedents were described across the 51 studies (Table 1).

**Table 1:** Summary of studies meeting criteria for data extraction.

|  | Country | Population | Decedents Included | Mean (Median) Age (years) | Number with autopsy IMD | Autopsy Type |
| --- | --- | --- | --- | --- | --- | --- |
| Beigmohammadi et al. (2020)<sup>20*</sup> | Iran | Hospitalized | 7† | 67·9 (72) | 0 | Minimally invasive |
| Borczuk et al. (2020)<sup>21</sup> | Italy, USA | Hospitalized | 68† | (73) | 2 | Standard; 2 minimally invasive |
| Böesmüller et al. (2020)<sup>22</sup> | Germany | Hospitalized | 4† | 71·8 (75) | 0 | Standard |
| Bradley et al. (2020)<sup>23*</sup> | USA | Hospitalized | 14† | 70·6 (73·5) | 0 | Standard |
| Brook et al. (2020)<sup>24</sup> | USA | Hospitalized | 5† | (79) | 0 | Minimally invasive |
| Bryce et al. (2020)<sup>25</sup> | USA | Hospitalized | 25 | ND | 0 | Standard |
| Buja et al. (2020)<sup>26</sup> | USA | Hospitalized | 3† | 48 (48) | 0 | Standard |
| Carsana et al. (2020)<sup>27*</sup> | Italy | Hospitalized | 38 | 69 | 1 | Standard |
| Copin et al. (2020)<sup>28*</sup> | France | Hospitalized | 6 | ND | 0 | Standard |
| De Michele et al. (2020)<sup>29*</sup> | USA | Hospitalized | 40 | (71·5) | 1 | Standard |
| Deinhardt-Emmer et al. (2020)<sup>30*</sup> | Germany | Hospitalized | 11† | 72·2 (78) | 1 | Standard |
| Dell'Aquila et al. (2020)<sup>31*</sup> | Italy | Hospitalized | 12 | 82·3 | 0 | Standard |
| Desai et al. (2020)<sup>32*</sup> | USA | Hospitalized | 20† | 62·5 | 0 | Standard |
| Duarte-Neto et al. (2020)<sup>33*</sup> | Brazil | Hospitalized | 10 | (69) | 0 | Minimally invasive |
| Elezkurtaj et al. (2020)<sup>34*</sup> | Germany | Hospitalized | 26† | 69 (69) | 1 | Standard |
| Elsoukkary et al. (2020)<sup>35</sup> | USA | Hospitalized | 9‡ | ND | 0 | Standard and minimally invasive |
| Falasca et al. (2020)<sup>36*</sup> | Italy | Hospitalized | 22† | 68 (69·5) | 0 | Standard |
| Flikweert et al. (2020)<sup>37</sup> | Netherlands | Hospitalized | 7† | (74) | 0 | Minimally Invasive |
| Fox et al. (2020)<sup>38</sup> | USA | Hospitalized | 10† | 63 (64·5) | 0 | Standard |
| Giacca et al. (2020)<sup>39</sup> | Italy | Hospitalized | 41† | 79·7 | 0 | Standard |
| Grosse et al. (2020)<sup>40</sup> | Austria | Hospitalized | 14† | 80·6 (81·5) | 0 | Standard |
| Hanley et al. (2020)<sup>41</sup> | United Kingdom | Hospitalized | 10† | (73) | 0 | Standard, 1 minimally invasive |
| Hellman et al. (2020)<sup>42*</sup> | Sweden | Hospitalized | 3 | ND | 0 | Standard |
| Kimmig et al. (2020)<sup>43*</sup> | USA | Hospitalized | 7 | ND | 0 | Standard |
| Kommos et al. (2020)<sup>44*</sup> | Germany | Hospitalized | 13† | 74·6 (78) | 0 | Standard |
| Konopka et al. (2020)<sup>45</sup> | USA | Hospitalized, Community | 8† | 55 (52) | 0 | Standard |
| Lax et al. (2020)<sup>46*</sup> | Austria | Hospitalized | 11† | 80·5 | 0 | Standard |
| Leppkes et al. (2020)<sup>47*</sup> | Germany | Hospitalized | 8 | ND | 0 | Standard |
| Li et al. (2020)<sup>48</sup> | China | Hospitalized | 30† | 68·2 (68·5) | 0 | Minimally invasive |
| Martines et al. (2020)<sup>49</sup> | USA | Hospitalized | 8† | 73·5 | 0 | Standard |
| Menter et al. (2020)<sup>50</sup> | Switzerland | Hospitalized | 21† | 76.4 (75) | 0 | Standard, some minimally invasive |
| Nagashima et al. (2020)<sup>51*</sup> | Brazil | ND | 6 | ND | 0 | Minimally invasive |
| Oprince and Muja. (2020)<sup>52</sup> | Romania | Hospitalized, Community | 3† | 58.7 (70) | 0 | Standard |
| Prieto-Perez et al. (2020)<sup>53*</sup> | Spain | Hospitalized | 20 | (79) | 0 | Minimally invasive |
| Prilutskiy et al (2020)<sup>54</sup> | USA | Hospitalized | 4† | 74 (72) | 0 | Standard |
| Radermecker et al (2020)<sup>55</sup> | Belgium | Hospitalized | 4† | 60.8 (59.5) | 0 | Standard |
| Rapkiewicz et al (2020)<sup>56*</sup> | USA | Hospitalized | 7† | 57.4 (60) | 1 | Standard |
| Rommelink et al (2020)<sup>57*</sup> | Belgium | Hospitalized | 17† | 67.5 (68) | 2 | Standard |
| Roden et al (2020)<sup>58</sup> | USA | Hospitalized | 8† | (79) | 0 | Standard |
| Roncati et al (2020)<sup>59*</sup> | Italy | Hospitalized | 3 | 53.3 (49) | 0 | Minimally invasive |
| Sadegh Beigee et al. (2020)<sup>60</sup> | Iran | Hospitalized | 25 | (66) | 0 | Minimally invasive |
| Sauter et al (2020)<sup>61</sup> | USA | Hospitalized | 1†‡ | ND | 0 | Standard |
| Schaefer et al (2020)<sup>62*</sup> | USA | Hospitalized | 7† | 62.4 (66) | 1 | Standard |
| Schaller et al (2020)<sup>63</sup> | Germany | Hospitalized | 10† | (79) | 0 | Standard |
| Schurink et al (2020)<sup>64*</sup> | Netherlands | Hospitalized | 18 | ND | 1 | Standard |
| Skok et al (2020)<sup>65</sup> | Austria | Hospitalized | 11‡ | ND | 0 | Standard |
| Sufang et al (2020)<sup>66*</sup> | China | Hospitalized | 4 | 73 (76) | 0 | Minimally invasive |
| Valdivia-Mazeyra et al (2020)<sup>67</sup> | Spain | Hospitalized | 18 | 61 | 0 | Standard; 7 minimally invasive |
| Wichmann et al (2020)<sup>68*</sup> | Germany | Hospitalized | 12† | (73) | 0 | Standard |
| Wu et al (2020)<sup>69*</sup> | Italy | Hospitalized | 10 | ND | 0 | Standard |
| Youd et al (2020)<sup>70</sup> | United Kingdom | Community | 3† | 82.3 (86) | 0 | Standard |

Legend: \* author contacted, † sufficient information for patient-level data extraction, ‡ some decedents included in other cohort therefore patient number reduced, ND not documented

Studies were included from 15 countries, most commonly from the USA (including 224 decedents), Italy (146) and Germany (84) (Figure 2). Most autopsies were performed on decedents who were hospitalized (n=694 [99%]) with very few from community (1%). Standard autopsy was the most commonly employed technique and was used in 36 (70·6%) studies. Minimally invasive was used in 10 (19·6%) studies and a combination of the two was used in five (9·8%) studies. Invasive mechanical ventilation and ECMO status was missing for 155 (22%) decedents. Of the total cohort, 314 (44·7%) required invasive mechanical ventilation and 15 (2·1%) were on ECMO.

**Figure 2.**
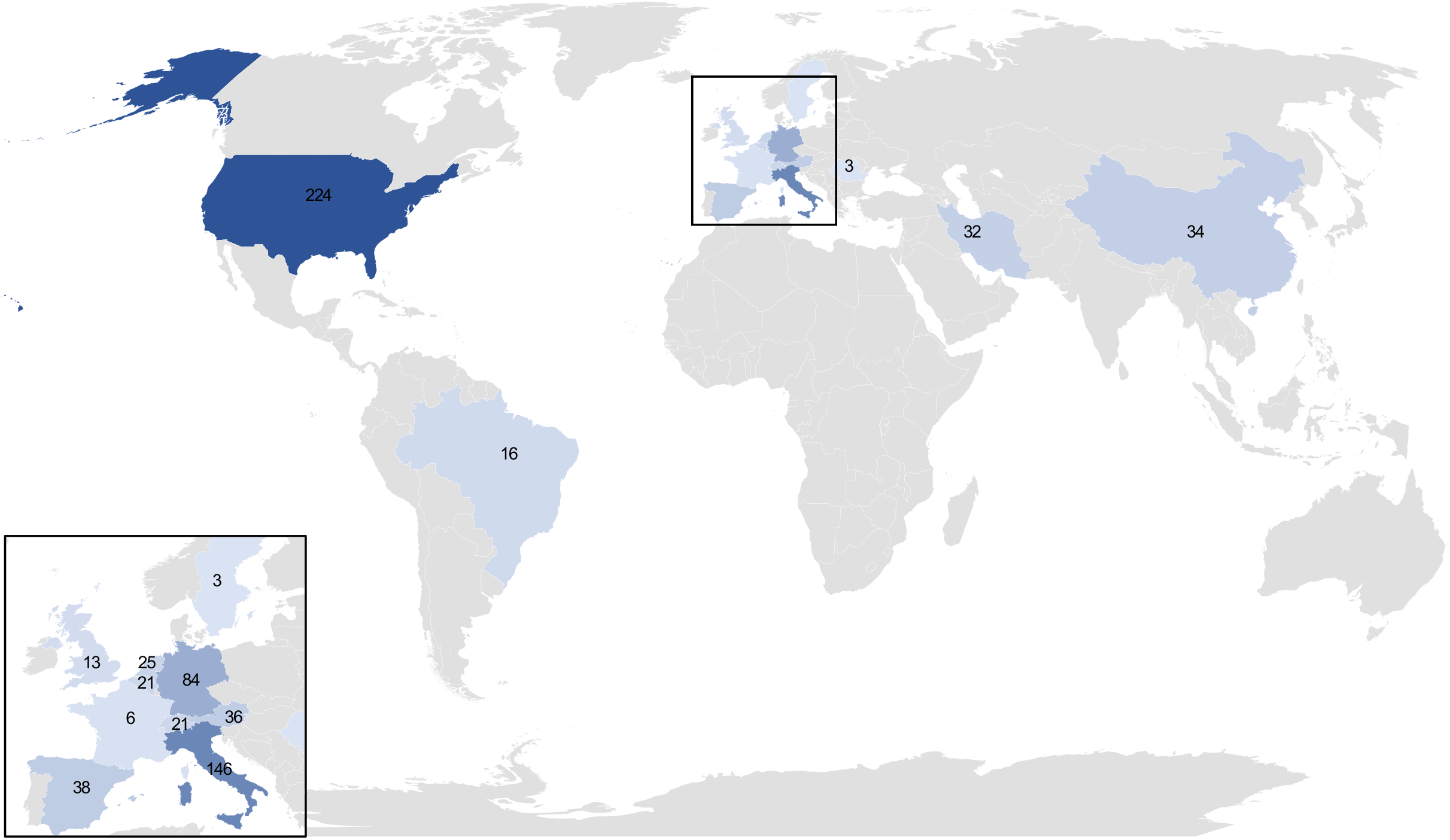

Enough information was provided for individual-level data extraction for 32 studies describing 430 decedents (Supplementary Table 1). The median age was 72 years (IQR 61 to 80 years). Most decedents had pre-existing comorbidities. Diabetes mellitus, pre-existing lung disease, and cancer, were reported for 129 (30%), 95 (22·1%), and 56 decedents (13%), respectively. Immunocompromised status was reported for 25 decedents (5.8%), including solid organ transplantation in nine (2·1%), hematologic malignancy in seven (1·6%), and chronic immunosuppressant use in nine (2·1%). The median duration of hospitalization before death was 10 days (IQR 5-22 days). Data on mechanical ventilation status was available for 335 decedents: 173 decedents (51·6%) had received mechanical ventilation for a median of nine days (IQR 5-20), whereas 162 (48·4%) died without mechanical ventilation. Treatment included immunomodulation for 60 decedents (seven received interleukin [IL]-6 inhibitors; two received IL-1 inhibitors; one received both of these drug classes; 48 received steroids; and details of immunomodulatory therapy missing for one). Forty-one decedents (9·5%) were recorded to have received antifungals.

Among the total cohort of 702 decedents, 11 (1·6%) had autopsy evidence of IMD, including six (0·9%) described as pulmonary aspergillosis, four (0·6%) with invasive mycoses not specified, and one (0·1%) with mucormycosis (Table 2). Among decedents with CAPA, disease was limited to the airways in one, involved only the lungs in four, and both airways and lung in one case. Of the decedents with unidentified IMD, disease was confined to the airways in one, and was found in the lungs in three. In the patient with mucormycosis, fungal invasion was demonstrated in lungs, hilar lymph nodes, brain, and kidney. Of 430 decedents with individual-level data, nine (2·1%) had autopsy-proven IMD including five (1·2%) with CAPA, three with unidentified invasive moulds (0·7%) and one with mucormycosis. Decedents with and without IMD that had patient-level data available are compared in Table 3.

**Table 2:** Details of decedents with IMD.

|  | Case number in report | Age, Gender | Length of illness (days) | Pre-existing immune-suppression | Immuno - therapy | IMV days | Antifungal | Species of mould | Type of autopsy | Extent of fungal involvement |
| --- | --- | --- | --- | --- | --- | --- | --- | --- | --- | --- |
| Borczuk et al.<sup>21</sup> | 29 | 79, M | 9 | None | None | 6 | None | *Aspergillus* | Standard; 2 in series minimally invasive | Airway only |
| Borczuk et al.<sup>21</sup> | 39 | 61, M | 6 | None | None | 0 | None | *Aspergillus* | Standard; 2 in series minimally invasive | Bronchopneumonia |
| Carsana et al.<sup>27</sup> | NA | NA | NA | NA | NA | NA | NA | Fungus not specified | Standard | Fungal pulmonary abscess |
| De Michele et al.<sup>29</sup> | NA | NA | NA | NA | NA | NA | NA | *Aspergillus* | Standard | Bronchopneumonia |
| Deinhardt-Emmer et al.<sup>30</sup> | 3 | 78, M | 30 | None | None | 7 | None | Fungus not specified | Standard | Fungal pneumonia |
| Elezkurtaj et al.<sup>34</sup> | 22 | 30, F | 21 | Prior allo-HSCT and GVHD | None | ND | ND | Fungus not specified | Standard | Invasive pulmonary mycosis |
| Hanley et al.<sup>41</sup> | 5 | 22, M | 27 | None | None | 22 | Caspofungin | Mucormycte | Standard | Lungs, hilar lymph nodes, brain, kidney |
| Rapkiewicz et al.<sup>56</sup> | 2 | 60, M | 7 | None | None | 0 | None | Fungus not specified | Standard | Erosive bronchitis with hyphae |
| Rommelink et al.<sup>57</sup> | 6 | 73, M | 11 | None | Steroids | Y, ND | ND | *Aspergillus* | Full lung biopsies | Lungs and airway |
| Rommelink et al.<sup>57</sup> | 7 | 56, M | 7 | None | None | 0 | ND | *Aspergillus* | Full lung biopsies | Lungs |
| Schaefer et al.<sup>62</sup> | 4 | 50, M | 9 | B-ALL | None | 7 | ND | *Aspergillus* * | Standard | Lung abscess |

Legend: \*pre-existing diagnosis, allo-HSCT allogeneic stem cell transplant, B-ALL b-cell acute lymphoblastic leukemia, GVHD graft versus host disease, IMV invasive mechanical ventilation, NA not applicable (not individual patient data in study), ND not documented

**Table 3:** Patient-level data for decedents with and without autopsy-proven invasive mould disease.

|  | Invasive mould disease<br>(n=9 [2.1%]) | No invasive mould disease<br>(n=421 [97.9%]) |
| --- | --- | --- |
| Age in years, median (IQR) | 60 (40-75.5) | 70 (57-79)* |
| Male | 8 (88.9%) | 260† (66.3%) |
| Pre-existing lung disease | 1 (11.1%) | 94 (22.3%) |
| Duration from symptom onset to death in days, median (IQR) | 9 (6.75-22.5)‡ | 14 (9-26)§ |
| Hospital death | 9 (100%) | 353 (83%) |
| Hospital LOS in days, median (IQR) | 16.5 (2.75-28.0)¶ | 10 (5-22)\|\| |
| Ventilated | 5 (55.5%)** | 168 (39.9%)†† |
| Length of ventilation in days, median (IQR) | 7 (6.25-18.25)‡‡ | 9 (4.75-9.0)§§ |
| Host-directed therapies for COVID-19 | 1 (11.1%) | 58 (13.7%) |

Legend: Data presented as n (%) unless otherwise specified. \*data missing for 60, † data missing for 29, ‡ data missing for 3, § data missing for 167, ¶ data missing for 5, || data missing for 112, \*\*data missing for 1, †† data missing for 94, ‡‡ data missing for 5, §§ data missing for 33

Two decedents with IMD (22·2%) had pre-existing immunocompromising conditions vs 23 (5·3%) without CAPA. In one immunocompromised decedent in the IMD group, the diagnosis of aspergillosis pre-dated COVID-19^62^ (Sanjat Kanjilal Brigham and Women’s Hospital, personal communication). Only one decedent (11·1%) with IMD received immunomodulatory therapy for COVID-19 compared to 59 (13·7%) without IMD. One decedent with IMD (11·1%) was recorded to have received antifungals.

## Discussion

In this systematic review of autopsy studies, we describe the prevalence of autopsy-proven IMD including CAPA among persons with fatal COVID-19 infections. IMD was autopsy-proven in just 1·6% of all decedents with COVID-19 and 2·9% among those reported to have received mechanical ventilation. While we cannot definitively conclude that the unidentified moulds seen on autopsy studies were *Aspergillus*, given that this is the most common cause of IMD, it is likely that most – if not all – were CAPA.

The rates of autopsy-proven IMD are considerably lower than rates of CAPA reported from ICU cohort studies. In a systematic review of ICU cohort studies, the median prevalence of CAPA was 21.1% (IQR 15.2-27.5%) (unpublished data). Given that post-mortem examinations are the most reliable way to confirm IMD, this review provides a more robust appraisal of the true prevalence of IMD complicating critical COVID-19 and suggests that the high CAPA rates published from cohort studies may be due to over-diagnosis. Indeed, previous autopsy studies have shown that pneumonias, in general, were over-called antemortem, and that discrepancy rates between clinical and necropsy findings were higher for respiratory tract infections than for other types of infection.^71^

CAPA may be over-diagnosed in ICU studies because of a lack of specificity in the classification of probable/putative and possible CAPA (which characterize the vast majority of cases reported to date). Indeed, there are several challenges that ICU studies face in determining the true prevalence of CAPA. Firstly, the clinical and radiographic signs of CAPA are difficult to distinguish from severe COVID-19 alone. Secondly, lower respiratory tract samples for diagnosing IPA are infrequently obtained in patients with COVID-19 because bronchoscopy is considered an aerosol generating procedure. Thirdly, the clinical definitions for CAPA are imperfect. Proven IPA – which relies on histological evidence of tissue invasion - is rarely diagnosed, and given the tenuous clinical status of most affected patients, biopsies are rarely performed. Established research criteria for the diagnosis of IPA, such as the updated definitions from the EORTC/MSGERC^17^ are of limited utility for diagnosing IPA in ICU patients because host factors are frequently absent.^18^ Moreover, classic radiographic findings seen in immunocompromised hosts are uncommon in patients without host factors. Alternative diagnostic criteria have been proposed and adapted for the study of influenza-associated pulmonary aspergillosis.^7,72^ Several definitions have been proposed for CAPA.^5,12,15^ However, in critically ill patients with COVID-19, radiographic findings of IMD are non-specific, serum galactomannan lacks sensitivity, and the presence of moulds in respiratory samples (detected by culture, PCR, or galactomannan) can represent airway colonization. That some patients with CAPA improve without antifungals argues against true invasive disease.^9^ Moreover, Flikweert et al reported that histological evidence of IPA was absent on post-mortem examination of six patients diagnosed with probable CAPA on the basis of bronchoalveolar lavage galactomannan (median optical density index, 4·35, IQR 3·63-4·40) and, in two, respiratory culture growing *A. fumigatus*.^37^ Taken together with our findings that IMD is uncommon on autopsy studies of decedents with COVID-19, these data suggest that most cases of probable/putative or possible CAPA may not represent true invasive disease.

Alternatively, CAPA has been associated with excess mortality.^11^ The clinical importance of *Aspergillus* in the airways of patients with critical COVID-19 is unclear, but the low prevalence of autopsy-proven IMD in such patients suggests that this finding may be a marker of COVID-19 severity and not a mediator of death. Nonetheless, clinical trials are underway to assess the effect of anti-mould fungal prophylaxis in critically ill COVID-19 patients.

CAPA has primarily been reported among critically ill COVID-19 patients, especially those requiring mechanical ventilation, but it unclear whether this reflects sampling bias (enhanced surveillance and access to lower respiratory samples in this patient population); confounding (*Aspergillus* in the airways and critical illness/ventilation are both associated with COVID-19 severity); or whether critical illness and mechanical ventilation truly confer increased risk of CAPA. In our study, all cases of COVID-19 were severe by definition, given that they resulted in death. However, only 45% of decedents had received mechanical ventilation. Five of eight decedents with autopsy-proven IMD (62·5%) for whom this data was available had received mechanical ventilation (compared to 168/327 without IMD [39·9%]). These data suggest that IMD (including CAPA) is not confined to mechanically ventilated patients and that higher rates in these patients may reflect a combination of disease severity and convenience of testing.

Immune dysregulation is implicated in the pathogenesis of severe COVID-19, and immunomodulatory therapies like corticosteroids and cytokine inhibitors (IL-1 and IL-6 inhibitors) have been widely used. Corticosteroid therapy is a known risk factor for IPA^17^ and IL-6 inhibitors could plausibly provoke IMD given impaired immune response to other pathogens.^73^ Only one (11·1%) of the nine decedents with IMD for whom treatment data was available had received these drugs, comparable to 14% in the non-IMD group. Importantly, most autopsies were conducted prior to the publication of the RECOVERY trial which showed dexamethasone improved mortality in patients with severe and critical COVID-19,^74^ and so corticosteroid prescribing may have increased since then. Further study will be needed to determine whether use of immunomodulatory therapy will be associated with tissue-proven IMD.

Post-mortem examination is the most definitive means available to detect IMD and is a particular strength of this study. The presence of hyphae on histopathologic investigation is simple to detect and it is therefore highly likely that the majority of these cases would have been identified. We also made every effort to contact authors when histopathologic findings were not specifically described to confirm that IMD had not been encountered in each case. Another strength is the inclusive nature of our review, which included pre-prints, given the rapidly expanding literature in COVID-19.

Despite these strengths, there are several limitations of this study that should be considered. First, we only included studies published in English or French, and it is possible that excluded studies could have contributed further data. Second, we were confronted with limited patient information from most reports. Use of antifungals was not commonly reported, and we cannot be sure whether this reflects incomplete data or low rates of prescribing. Antifungal use could have reduced the sensitivity of IMD detection on autopsy. The use of immunomodulators was also incompletely reported. While this would be unlikely to affect the main findings, this data could be helpful in the understanding of risk of tissue-proven IMD associated with these therapies. Third, although our review was exhaustive, the sample size is very small compared the total number of COVID-19 deaths internationally; most decedents were included from just three countries (US, Germany, Italy) and geographic variation in incidence of IMD following COVID-19 may occur (as it does following influenza).^75^ Fourth, while the use of minimally invasive autopsy strategies for COVID-19 have been validated,^25,33,76^ random sampling rather than whole lung examination may have led to under-diagnosis of IMD; it is re-assuring, however, that the majority of our included cases were performed with standard technique.

In conclusion, autopsy-proven IMD is uncommonly reported among decedents with COVID-19. In order to further our understanding of the prevalence of IMD, future autopsy studies should include patients with ante-mortem diagnoses of CAPA to validate clinical definitions for this disease. Future autopsy studies should specifically consider co-infections and make note of clinical diagnoses and antemortem use of antimicrobials, including antifungals, and of immunomodulatory treatments.

## Panel – Research in Context

### Evidence before this study

We searched 4 databases dated from January 1, 2019 to September 26, 2020 for studies written in English or French describing lung histopathology in autopsy specimens from decedents with COVID-19, with the view of outlining the prevalence of tissue-invasive mould infection. Estimating the prevalence of CAPA in clinical studies has been limited by scarcity of biopsies in critically ill patients, and reliance on various proposed definitions that combine non-specific clinical, radiographic, and microbiological findings to make diagnoses of probable (putative) or possible CAPA. There are currently no published systematic reviews focusing on COVID-19 autopsy cases for secondary IMD, including CAPA, which would be needed to meet definitive criteria.

### Added value of the study

We did a systematic review of autopsy series including ≥3 decedents diagnosed pre- or post-mortem with COVID-19 and identified 51 studies that met inclusion criteria. Data was from 16 countries and identified 702 decedents. Given that tissue sampling is the gold standard for diagnosis of IMD, this is the best available evidence to support that IMD including CAPA is rare in COVID-19 patients. This was far less common (1·7%) than that reported in clinical case series. There was inconsistent reporting to review potential risk factors including immunocompromised status, invasive mechanical ventilation, and exposure to immunomodulatory therapy.

### Implications of all the available evidence

Our findings suggest that tissue-invasive IMD including CAPA and complicating COVID-19 is rare. The presence of aspergillosis on culture is not diagnostic of clinical infection and may represent colonization. Further autopsy studies specifically directed at fungal super-infections would be optimal to further delineate the true frequency of infection.

## Data Availability

All relevant data included in supplemental table

## Contributors

BEK conducted the literature search and wrote the first draft of the manuscript. BEK and ISS did the review, data extraction and analysis, and created the figures. ISS was responsible for study design and overall supervision. All authors contributed to the data interpretation, and to critical revision of the manuscript.

## References

1. Maiese A, Manetti AC, La Russa R, Di Paolo M, Turillazzi E, Frati P, et al. Autopsy findings in COVID-19-related deaths: A literature review. Forensic Sci Med Pathol 2020; published online Oct 7. DOI10.1007/s12024-020-00310-8.

2. Thompson Iii GR, Cornely OA, Pappas PG, et al. Invasive aspergillosis as an under-recognized superinfection in COVID-19. Open Forum Infect Dis 2020; 7: ofaa242.

3. Hoenigl M. Invasive fungal disease complicating COVID-19: When it rains it pours. Clin Infect Dis 2020; published online Sep 5. DOI 10.1093/cid/ciaa1342.

4. Armstrong-James D, Youngs J, Bicanic T, Abdolrasouli A, et al. Confronting and mitigating the risk of COVID-19 associated pulmonary aspergillosis. Eur Respir J 2020; 56: 2002554.

5. Verweij PE, Gangneux JP, Bassetti M, et al. Diagnosing COVID-19-associated pulmonary aspergillosis. Lancet Microbe 2020; 1: e53–e55.

6. Rijnders BJA, Schauwvlieghe, A. F. A. D., Wauters J. Influenza-associated pulmonary aspergillosis: A local or global lethal combination? Clin Infect Dis 2020; 71: 1764–1767.

7. Schauwvlieghe, A. F. A. D., Rijnders BJA, Philips N, et al. Invasive aspergillosis in patients admitted to the intensive care unit with severe influenza: A retrospective cohort study. Lancet Respir Med 2018; 6: 782–792.

8. Lahmer T. Invasive pulmonary aspergillosis in critically ill patients with severe COVID-19 pneumonia: Results from the prospective AspCOVID-19 study. medRxiv 2020. DOI:2020.07.21.20158972.

9. Alanio A, Dellière S, Fodil S, Bretagne S, Mégarbane B. Prevalence of putative invasive pulmonary aspergillosis in critically ill patients with COVID-19. Lancet Respir Med 2020; 8: e48–e49.

10. Rutsaert L, Steinfort N, Van Hunsel T, et al. COVID-19-associated invasive pulmonary aspergillosis. Ann Intensive Care 2020; 10: 71–74.

11. Bartoletti M, Pascale R, Cricca M, et al. Epidemiology of invasive pulmonary aspergillosis among intubated patients with COVID-19: A prospective study. Clin Infect Dis 2020; published online Jul 28. DOI:10.1093/cid/ciaa1065..

12. White PL, Dhillon R, Cordey A, et al. A national strategy to diagnose coronavirus disease 2019–Associated invasive fungal disease in the intensive care unit. Clin Infect Dis 2020; published online Aug 29. DOI:10.1093/cid/ciaa1298.

13. Dupont D, Menotti J, Turc J, et al. Pulmonary aspergillosis in critically ill patients with coronavirus disease 2019 (COVID-19). Med Mycol 2020; published online Sep 10. DOI:10.1093/mmy/myaa078.

14. Nasir N, Farooqi J, Mahmood SF, Jabeen K. COVID-19-associated pulmonary aspergillosis (CAPA) in patients admitted with severe COVID-19 pneumonia: An observational study from Pakistan. Mycoses 2020; 63: 766–770.

15. Koehler P, Bassetti M, Chakrabarti A, et al. Defining and managing COVID-19 associated pulmonary aspergillosis: The 2020 ECMM/ISHAM consensus for research and clinical guidance. Lancet Infect Dis 2020; In Press.

16. Brown LK, Ellis J, Gorton R, De S, Stone N. Surveillance for COVID-19-associated pulmonary aspergillosis. Lancet Microbe 2020; 1: e152.

17. Donnelly JP, Chen SC, Kauffman CA, et al. Revision and update of the consensus definitions of invasive fungal disease from the european organization for research and treatment of cancer and the mycoses study group education and research consortium. Clin Infect Dis 2019; 71: 1367–1376.

18. Blot SI, Taccone FS, Van den Abeele, et al. A clinical algorithm to diagnose invasive pulmonary aspergillosis in critically ill patients. Am J Respir Crit Care Med 2012; 186: 56–64.

19. Liberati A, Altman DG, Tetzlaff J, et al. The PRISMA statement for reporting systematic reviews and meta-analyses of studies that evaluate healthcare interventions: explanation and elaboration. BM J 2009; 21: b2700.

20. Beigmohammadi MT, Jahanbin B, Safaei M, et al. Pathological findings of postmortem biopsies from lung, heart, and liver of 7 deceased COVID-19 patients. Int j Surg Path 2020; published online Jun 19. DOI:10.1177/1066896920935195.

21. Borczuk AC, Salvatore SP, Seshan S,V, et al. COVID-19 pulmonary pathology: A multi-institutional autopsy cohort from Italy and New York City. Mod Path 2020; 33: 2156–2168.

22. Boesmueller H, Traxler S, Bitzer M, et al. The evolution of pulmonary pathology in fatal COVID-19 disease: An autopsy study with clinical correlation. Virchows Arch 2020; 477: 349–357.

23. Bradley BT, Maioli H, Johnston R, et al. Histopathology and ultrastructural findings of fatal COVID-19 infections in washington state: A case series. Lancet 2020; 396: 320–332.

24. Brook OR, Piper KG, Mercado NB, et al. Feasibility and safety of ultrasound-guided minimally invasive autopsy in COVID-19 patients. Abdom Radiol (NY) 2020; published online Sep17. DOI:10.1007/s00261-020-025753-7.

25. Bryce C, Grimes Z, Pujadas E, et al. Pathophysiology of SARS-CoV-2: Targeting of endothelial cells renders a complex disease with thrombotic microangiopathy and aberrant immune response. The Mount Sinai COVID-19 autopsy experience. medRxiv 2020. DOI:10.1101/2020.05.18.20099960.

26. Buja LM, Wolf DA, Zhao B, et al. The emerging spectrum of cardiopulmonary pathology of the coronavirus disease 2019 (COVID-19): Report of 3 autopsies from Houston, Texas, and review of autopsy findings from other United States cities. Cardiovasc Pathol 2020; 48: 107233.

27. Carsana L, Sonzogni A, Nasr A, et al. Pulmonary post-mortem findings in a series of COVID-19 cases from northern Italy: A two-centre descriptive study. Lancet Infect Dis 2020; 20: 1135–1140.

28. Copin M, Parmentier E, Duburcq T, et al. Time to consider histologic pattern of lung injury to treat critically ill patients with COVID-19 infection. Intensive Care Med 2020; 46: 1124–1126.

29. De Michele S, Sun Y, Yilmaz MM, Katsyv I, Salvatore M, Dzierba AL, et al. Forty postmortem examinations in COVID-19 patients. Am J Clin Pathol 2020; 154: 748–760.

30. Deinhardt-Emmer S, Wittschieber D, Sanft J, et al. Early postmortem mapping of SARS-CoV-2 RNA in patients with COVID-19 and correlation to tissue damage. bioRxiv 2020. DOI:10.1101/2020.07.01.182550.

31. Dell’Aquila M, Cattani P, Fantoni M, et al. Postmortem swabs in the sars-CoV-2 pandemic: Report on 12 complete clinical autopsy cases. Arch Pathol Lab Med 2020; 144: 1298–1302.

32. Desai N, Neyaz A, Szabolcs A, et al. Temporal and spatial heterogeneity of host response to SARS-CoV-2 pulmonary infection. medRxiv 2020. DOI:10.1101/2020.07.30.20165241.

33. Duarte-Neto AN, de Almeida Monteiro, R. A., da Silva, et al. Pulmonary and systemic involvement of COVID-19 assessed by ultrasound-guided minimally invasive autopsy. Histopathology 2020; 77: 186–197.

34. Elezkurtaj S, Greuel S, Ihlow J, et al. Causes of death and comorbidities in patients with COVID-19. medRxiv 2020. DOI:10.1101/2020.06.15.20131540.

35. Elsoukkary SS, Mostyka M, Dillard A, et al. Autopsy findings in 32 patients with COVID-19: A single-institution experience. Pathobiology 2020; published online Sep 17. DOI:10.1159/000511325.

36. Falasca L, Nardacci R, Colombo D, et al. Post-mortem findings in italian patients with COVID-19 - a descriptive full autopsy study of cases with and without co-morbidities. J Infect Dis 2020; 222: 1807–1815.

37. Flikweert AW, Grootenboers, Marco J. J. H., et al. Late histopathologic characteristics of critically ill COVID-19 patients: Different phenotypes without evidence of invasive aspergillosis, a case series. J Crit Care 2020; 59: 149–155.

38. Fox SE, Akmatbekov A, Harbert JL, Li G, Brown JQ, Heide RSV. Pulmonary and cardiac pathology in African American patients with COVID-19: An autopsy series from New Orleans. Lancet Respir Med 2020; 8: 681–686.

39. Giacca M, Bussani R, Schneider E, et al. Persistence of viral RNA, widespread thrombosis and abnormal cellular syncytia are hallmarks of COVID-19 lung pathology. medRxiv 2020. DOI:10.1101/2020.06.22.20136358.

40. Grosse C, Grosse A, Salzer HJF, Dunser MW, Motz R, Langer R. Analysis of cardiopulmonary findings in COVID-19 fatalities: High incidence of pulmonary artery thrombi and acute suppurative bronchopneumonia. Cardiovasc Pathol 2020; 49: 107263.

41. Hanley B, Naresh KN, Roufosse C, et al. Histopathological findings and viral tropism in UK patients with severe fatal COVID-19: A post-mortem study. Lancet Microbe 2020; 1: e245–253.

42. Hellman U, Karlsson MG, Engström-Laurent A, et al. Presence of hyaluronan in lung alveoli in severe Covid-19 - an opening for new treatment options? J Biol Chem 2020; 295: 15418–15422.

43. Kimmig LM, Wu D, Gold M, et al. IL6 inhibition in critically ill COVID-19 patients is associated with increased secondary infections. Front Med (Lausanne) 2020; 7: 583897.

44. Kommoss FKF, Schwab C, Tavernar L, et al. The pathology of severe COVID-19-related lung damage. Dtsch Arztebl Int 2020; 117: 500–506.

45. Konopka KE, Nguyen T, Jentzen JM, et al. Diffuse alveolar damage (DAD) resulting from coronavirus disease 2019 infection is morphologically indistinguishable from other causes of DAD. Histopathology 2020; 77: 570–578.

46. Lax SF, Skok K, Zechner P, et al. Pulmonary arterial thrombosis in COVID-19 with fatal outcome results from a prospective, single-center, clinicopathologic case series. Ann Intern Med 2020; 173: 350–361.

47. Leppkes M, Knopl J, Naschberger E, et al. Vascular occlusion by neutrophil extracellular traps in COVID-19. Ebiomedicine 2020; 58: 102925.

48. Li Y, Wu J, Wang S, et al. Progression to fibrosing diffuse alveolar damage in a series of 30 minimally invasive autopsies with COVID-19 pneumonia in Wuhan, China. Histopathology 2020; published online Sep 14. DOI:10.1111/his.14249.

49. Martines R, Ritter J, Matkovic E, et al. Pathology and pathogenesis of SARS-CoV-2 associated with fatal coronavirus disease, United States. Emerg Infect Dis 2020; 26: 2005–2015.

50. Menter T, Haslbauer JD, Nienhold R, et al. Postmortem examination of COVID-19 patients reveals diffuse alveolar damage with severe capillary congestion and variegated findings in lungs and other organs suggesting vascular dysfunction. Histopathology 2020; 77: 198–209.

51. Nagashima S, Mendes MC, Camargo Martins AP, et al. Endothelial dysfunction and thrombosis in patients with COVID-19-brief report. Arterioscler Thromb Vasc Biol 2020; 40: 2404–2407.

52. Oprinca G, Muja L. Postmortem examination of three SARS-CoV-2-positive autopsies including histopathologic and immunohistochemical analysis. Int J Legal Med 2020; published online Aug 27. DOI:10.1007/s00414-020-02406-w.

53. Prieto-Perez L, Fortes J, Soto C, et al. Histiocytic hyperplasia with hemophagocytosis and acute alveolar damage in COVID-19 infection. Mod Pathol 2020; 33: 2139–2146.

54. Prilutskiy A, Kritselis M, Shevtsov A, et al. SARS-CoV-2 infection-associated hemophagocytic lymphohistiocytosis. Am J Clin Pathol 2020; 154: 466–474.

55. Radermecker C, Detrembleur N, Guiot J et al. Neutrophil extracellular traps infiltrate the lung airway, interstitial, and vascular compartments in severe COVID-19. J Exp Med 2020; 217: e20201012.

56. Rapkiewicz AV, Mai X, Carsons SE, et al. Megakaryocytes and platelet-fibrin thrombi characterize multi-organ thrombosis at autopsy in COVID-19: A case series. EClinicalMedicine 2020; 24: 100434.

57. Remmelink M, De Mendonca R, D’Haene N, et al. Unspecific post-mortem findings despite multiorgan viral spread in COVID-19 patients. Crit Care 2020; 24: 495.

58. Roden AC, Bois MC, Johnson TF, et al. The spectrum of histopathologic findings in lungs of patients with fatal COVID-19 infection. Arch Pathol Lab Med 2020; published online Aug 21. DOI:10.5858/arpa.2021-0491-SA.

59. Roncati L, Ligabue G, Nasillo V, et al. A proof of evidence supporting abnormal immunothrombosis in severe COVID-19: Naked megakaryocyte nuclei increase in the bone marrow and lungs of critically ill patients. Platelets 2020; 31: 1085–1089.

60. Sadegh Beigee F, Pourabdollah Toutkaboni M, et al. Diffuse alveolar damage and thrombotic microangiopathy are the main histopathological findings in lung tissue biopsy samples of COVID-19 patients. Pathol Res Pract 2020; 216: 153228.

61. Sauter JL, Baine MK, Butnor KJ, et al. Insights into pathogenesis of fatal COVID-19 pneumonia from histopathology with immunohistochemical and viral RNA studies. Histopathology 2020; 77: 915–925.

62. Schaefer I, Padera RF, Solomon IH, et al. In situ detection of SARS-CoV-2 in lungs and airways of patients with COVID-19. Mod Pathol 2020; 33: 2104–2114.

63. Schaller T, Hirschbuhl K, Burkhardt K, et al. Postmortem examination of patients with COVID-19. JAMA 2020; 323: 2518–2520.

64. Schurink B, Roos E, Radonic T, et al. Viral presence and immunopathology in patients with lethal COVID-19: A prospective autopsy cohort study. Lancet Microbe 2020; 1: e290–e299.

65. Skok K, Stelzl E, Trauner M, Kessler HH, Lax SF. Post-mortem viral dynamics and tropism in COVID-19 patients in correlation with organ damage. Virchows Arch 2020; published online Aug 20. DOI:10.1007/s00428-020-02903-8.

66. Sufang T, Yong X, Huan L, et al. Pathological study of the 2019 novel coronavirus disease (COVID-19) through postmortem core biopsies. Mod Pathol 2020; 33: 1007–1014.

67. Valdivia-Mazeyra M, Salas C, Nieves-Alonso J, et al. Increased number of pulmonary megakaryocytes in COVID-19 patients with diffuse alveolar damage: An autopsy study with clinical correlation and review of the literature. Virchows Arch 2020; published online Sep 11. DOI:10.1007/s00428-020-02926-1.

68. Wichmann D, Sperhake J, Luegehetmann M, et al. Autopsy findings and venous thromboembolism in patients with COVID-19. Ann Intern Med 2020; 173: 268–277.

69. Wu MA, Fossali T, Pandolfi L, et al. COVID-19: The key role of pulmonary capillary leakage. an observational cohort study. medRxiv 2020. DOI:10.1101/2020.05.17.20104877.

70. Youd E, Moore L. COVID-19 autopsy in people who died in community settings: The first series. J Clin Pathol 2020; 73: 840–844.

71. Gibson TN, Shirley SE, Escoffery CT, Reid M. Discrepancies between clinical and postmortem diagnoses in Jamaica: A study from the university hospital of the West Indies. J Clin Pathol 2004; 57: 980–985.

72. Verweij PE, Rijnders BJA, Brüggemann RJM, et al. Review of influenza-associated pulmonary aspergillosis in ICU patients and proposal for a case definition: An expert opinion. Intensive Care Med 2020; 46: 1524–1535.

73. Ching CB, Gupta S, Li B, et al. Interleukin-6/Stat3 signaling has an essential role in the host antimicrobial response to urinary tract infection. Kidney Int 2018 ; 93: 1320–1329.

74. RECOVERY Collaborative Group, Horby P, Lim WS, Emberson JR, et al. Dexamethasone in hospitalized patients with Covid-19 - preliminary report. N Engl J Med 2020; published online Jul 17. DOI:10.1056/NEJMoa2021436.

75. Schwartz IS, Friedman DZP, Zapernick L, et al. High rates of influenza-associated invasive pulmonary aspergillosis may not be universal: A retrospective cohort study from Alberta, Canada. Clin Infect Dis 2020; 71: 1760–1763.

76. Monteiro RAA, Duarte-Neto AN, Silva, L. F. F. D., et al. Ultrasound-guided minimally invasive autopsies: A protocol for the study of pulmonary and systemic involvement of COVID-19. Clinics (Sao Paulo) 2020; 75: e1972.

